# Impact of vaccination in reducing Hospital expenses, Mortality and Average length of stay among COVID-19 patients – a retrospective cohort study from India

**DOI:** 10.1101/2021.06.18.21258798

**Authors:** Madhumathi Ramakrishnan, Prakash Subbarayan

**Affiliations:** Star Health Insurance & Allied Limited, Chennai, India

**Author notes:** **Corresponding author**: Dr. Madhumathi Ramakrishnan, 3/5 Jai Nagar 7^th^ street, Arumbakkam, Chennai, **Email:**. **Funding:** Nil. **Ethical clearance:** Institutional Ethics Committee of Star Health & allied Insurance, Chennai approved the study on 01/Feb/2021 with an advice to maintain patient confidentiality at all stages.

**Keywords:** COVID-19, vaccination, hospital expenses, average length of stay

## Abstract

**Background & Aim:** WHO listed vaccine hesitancy among the top 10 global threats to health and there are very few reports highlighting vaccine benefits against COVID-19. The aim of this study was to study the impact of vaccination on reducing the average length of stay (ALOS), intensive care unit (ICU) requirement, mortality and cost of the treatment among COVID-19 patients.

**Methods:** In this retrospective cohort study all the patients above 45 years who underwent treatment for COVID-19 were included. The data of patients treated pan India during the period March & April 2021 with the diagnosis of COVID-19, under health insurance cover, were extracted to study parameters like the ALOS, mortality, ICU requirement, total hospital expenses incurred and the vaccination status.

**Results:** Among 3820 patients with COVID-19, 3301 (86.4%) were unvaccinated while 519 (13.6%) were vaccinated. Among the unvaccinated the mean (s.d) ALOS was 7 days. Fourteen days after second dose of vaccination this was significantly less (p=0.01) at 4.9. The mean total hospital expense among the unvaccinated was Rs. 277850. Fourteen days after second dose of vaccination this was further less (p=0.001) at Rs. 217850. Among the unvaccinated population 291/3301 (8.8%) required ICU and this was significantly less (p=0.03) at 31/519 (6%) among the vaccinated. Among those who received two doses of vaccination it was further less at 1/33 (3%). The mortality among unvaccinated patients was 16/3301 (0.5%) while there was no mortality among the vaccinated. Among those who received two doses of vaccination there was a 66% relative risk reduction in ICU stay and 81% relative risk reduction in mortality.

**Conclusions:** There was a significant reduction in ALOS, ICU requirement, mortality & treatment cost in patients who had completed two doses of vaccination. These findings may be used in motivating public and promoting vaccination drive.

## Introduction

Discovery of vaccination against Covid-19 is considered as one of the fastest vaccine research that has happened so far. It was rolled out in India for health care workers from January 16^th^ 2021, senior citizens from February2021, and for those above 45 years from March2021. While there are many publications from the western world regarding efficacy of vaccines, there are very few reports of vaccine efficacy from the Indian subcontinent.

Two different types of vaccines are used in the Indian setting. The first one to get approval was Oxford–AstraZeneca chimpanzee adenovirus vector vaccine ChAdOx1 nCoV-19 (AZD1222) which been studied extensively with many reports supporting its benefits [1,2]. The second one to be rolled out on trial basis was BBV152 whole-virion inactivated vero cell-derived vaccine [3]. The National Vaccination Bureau reported that 0.03-0.04% of persons who have received vaccination could still contract Covid-19(breakthrough infection), although the mortality and ICU stay reduced in them[4]. Further studies from the UK, have shown that even a single dose of vaccination may have a good protective effect[5]. The aim of this study was to study the impact of vaccination on reducing the average length of stay (ALOS), intensive care unit (ICU) requirement, mortality and cost of the treatment among COVID-19 patients.

## Methods

In this retrospective cohort study all the patients above 45 years who underwent treatment for COVID-19 were included. The patients aged 45 years and below were excluded as the vaccination program had not commenced for this population in India, during the study period. The data of patients treated pan India during the period March & April 2021 with the diagnosis of COVID-19, under health insurance cover, were extracted from Star Health Insurance, claims data-base using the ICD (U07.1 & U07.2) codes. Institutional ethical clearance was obtained and patient confidentiality was maintained with utmost care.

Study parameters like the Average length of stay (ALOS), mortality, intensive care unit (ICU) requirement, total hospital expenses and the vaccination status (first or second dose, time lapse in contracting disease among the vaccinated; and reasons for not vaccinating among the rest)were procured from the case records submitted. Wherever further information was required, the patients or their relatives were contacted through a telephonic call. The parameters were compared between the unvaccinated and vaccinated groups. Results were expressed as percentage, mean (s.d) and compared using chi square test or student’s t-test using graph-pad prism software and the difference was considered significant if the p value was less than 0.05.

## Results

A total of 3820 patients with COVID-19 were analyzed. Among this group, 3301 (86.4%) were unvaccinated while 519 (13.6%) were vaccinated. A total of 486 (12.7%) patients had received first dose, while 33 (0.8%) had received second dose. In 43% of the unvaccinated group there was no valid reason for the same. Table 1 describes the other reasons among the unvaccinated (apprehension/fear 21%, ignorance 14%; recent infections 9%; social factors 7% and lack of interest 6%).

**Table 1.** Reasons for not taking COVID vaccine (n=3301)

|  |  |  |
| --- | --- | --- |
| No valid reason | 1420 | 43% |
| Apprehension/Fear | 693 | 21% |
| Ignorance regarding age eligibility | 462 | 14% |
| Recent other infections | 297 | 9% |
| Family commitment/ Social factors | 231 | 7% |
| Disinterested | 198 | 6% |

Figure 1a describes the ALOS and hospital expense. Among the unvaccinated the mean (s.d) ALOS was 7 days. Fourteen days after second dose of vaccination this was significantly less (p=0.01) at 4.9 days. The mean total hospital expense among the unvaccinated was Rs. 277850. Fourteen days after second doses of vaccination this was further less (p=0.001) at Rs. 217850 (figure 1b).

**Figure 1.**
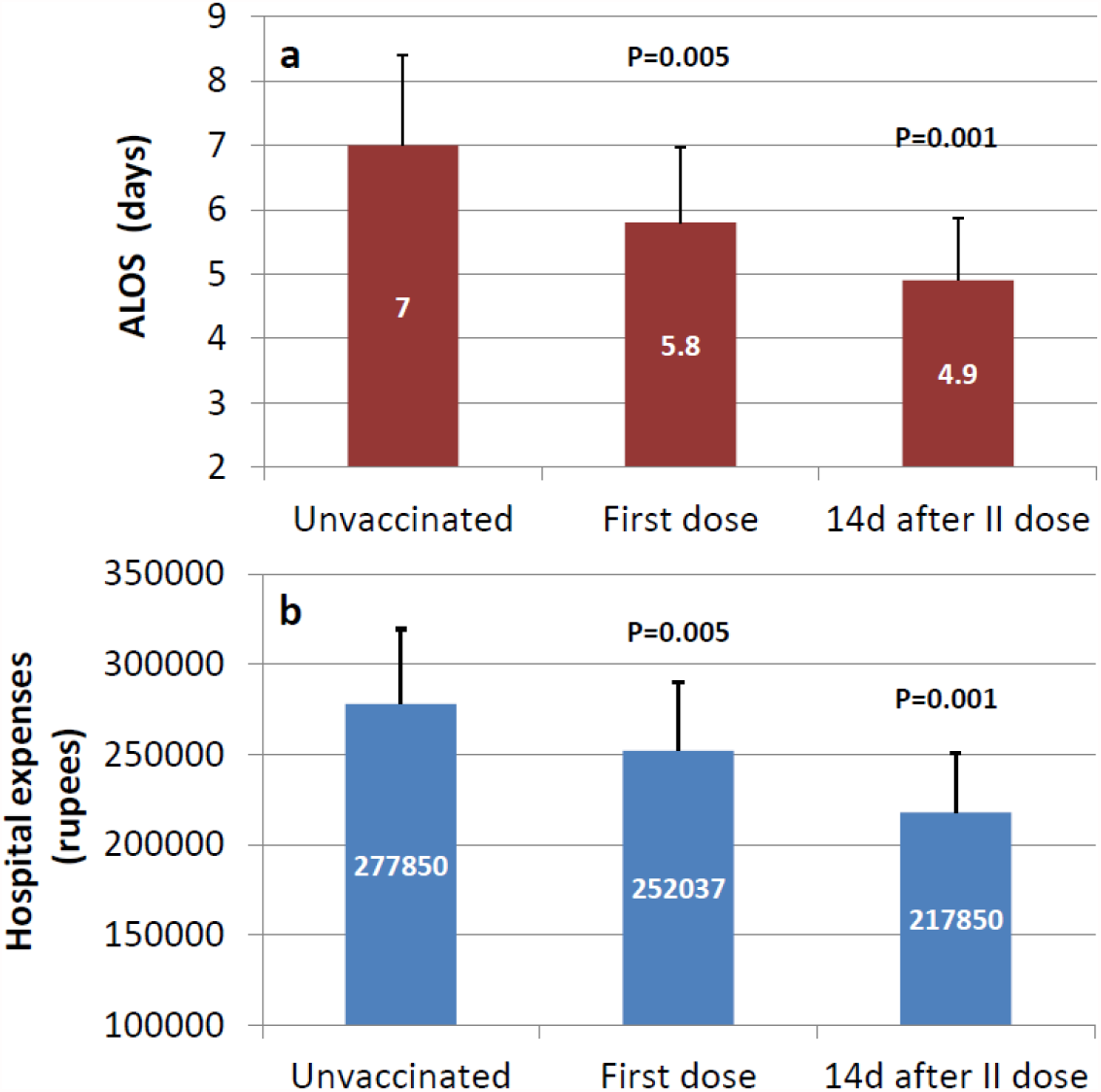
Comparison of a) Average length of stay (ALOS) and b) hospital expenses between unvaccinated versus those who completed first/ second dose of vaccination

Figure 2 compares ICU requirement between the groups. Among the unvaccinated population 291/3301 (8.8%) required ICU stay and this was significantly less (p=0.03) at 31/519 (6%) among the vaccinated. Among those who received two doses of vaccination it was further less at 1/33 (3%).The relative risk of ICU stay among unvaccinated population was 0.34 (95% C.I 0.49 – 2.37) and there was a 66% relative risk reduction in ICU stay after two doses of vaccination.

**Figure 2.**
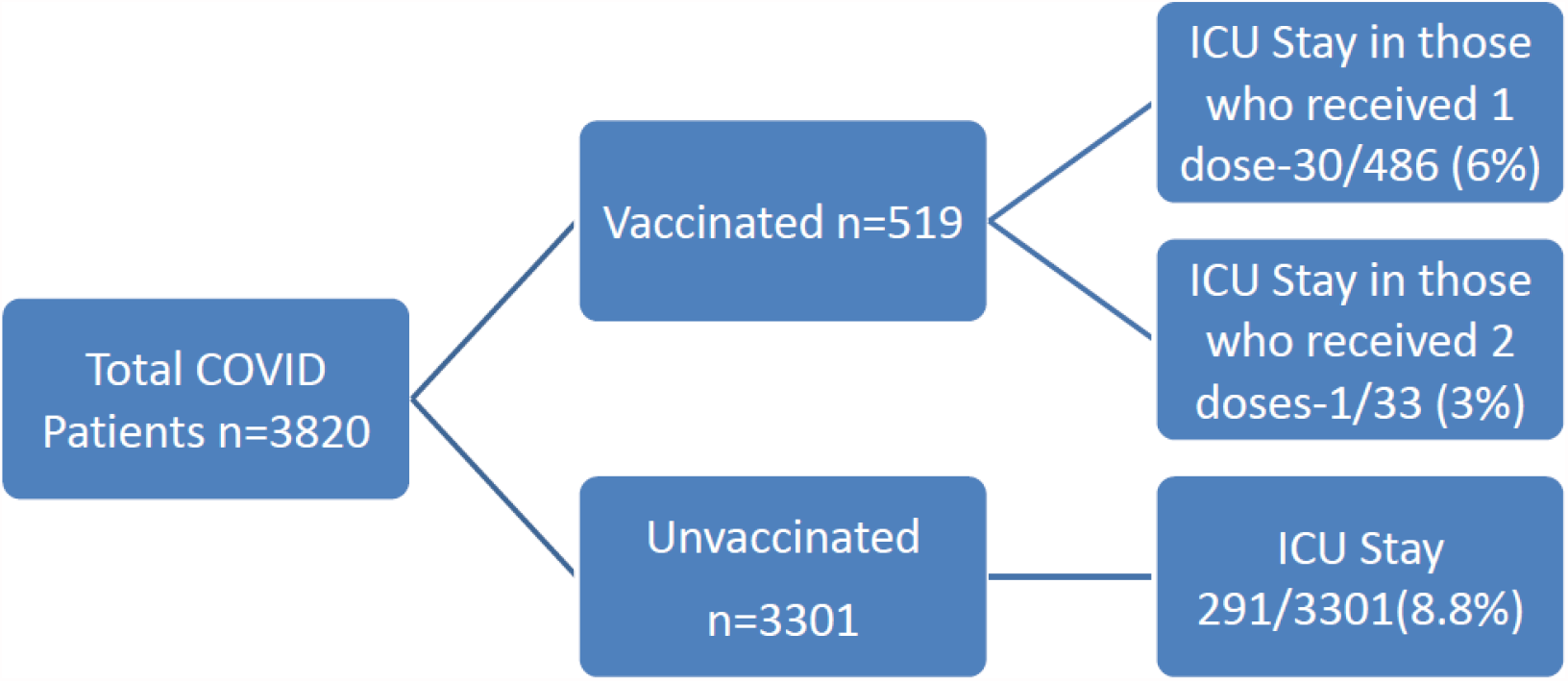
ICU stay based on vaccination status

The mortality among unvaccinated patients was 16/3301 (0.5%) while there was no mortality among the vaccinated patients (table 2). The relative risk of mortality among unvaccinated population was 0.19 (95% CI 0.02-3.20) and there was 81% relative risk reduction in mortality by vaccination.

**Table 2.** Comparison of ALOS, cost and ICU stay between vaccinated & Un-vaccinated.

| **Total Surveyed:3820** | **Un-vaccinated** | **Vaccinated** | **Significance**<br>**(p value)** |
| --- | --- | --- | --- |
|  | **(n=3301)** | **(n=519)** |  |
| Mean (s.d) Age | 54.1 (6.5) | 59.3 (5.8) | 0.001 |
| Male: Female: | 2182/1119 | 346/173 | 0.6 |
| ALOS in days | 7 (3.07) | 5.7 (3) | 0.001 |
| Treatment cost in rupees;<br>mean (s.d) | 277850 (41573) | 250116(32432) | 0.001 |
| No of patients needing ICU stay | 291 (8.8%) | 31 (6%) | 0.03 |
| Number of Deaths | 16 (0.5%) | 0 | n/a |

Table 3 sub classifies the outcomes among those with co-morbidities. Among the vaccinated patients, co-morbidities were found in 239/519 (46%) and this was significantly higher (p=0.001) than 1177/3301 (36%) noted in the un-vaccinated group. Despite having higher co-morbidities ALOS, ICU requirement and hospital expenses were significantly less (table 3) among the vaccinated.

**Table 3.** Co-morbidity and outcomes.

| **Comorbid conditions** | **Un-vaccinated**<br>**(n=3301)** | **Vaccinated**<br>**(n=519)** | **Significance**<br>**(p value)** |
| --- | --- | --- | --- |
| DM/HTN or both | 1177/3301(36%) | 239/519 (46%) | 0.001 |
| ALOS | 7.7 (3.06) | 5.9 (3.04) | 0.001 |
| Treatment cost in rupees<br>mean (s.d) | 294421 (30697) | 251302(31176) | 0.001 |
| ICU required | 111/1177 (9.4%) | 12/239 (5%) | 0.03 |

## Discussion

Breakthrough in vaccine discovery happened at lightning speed for COVID-19 compared to all other pandemics in the history, although the roll-out has been marred by multiple factors like availability, logistics, population size etc. In addition, the concerns on efficacy, risks and lack of solid trial data during the initial phases made people skeptical about vaccination. Although the importance of race against time to vaccinate public have been stressed by several authors [6,7] many countries including India could not fast track vaccination and encountered a huge second wave. While 0.03-0.04% of those who are vaccinated can still contract COVID-19 infection, vaccination is known to reduce hospital stay and mortality in them [8,9]. Thompson et al [10] reported 0.04 infections per 1,000 person-days among those fully immunized with m-RNA vaccine (≥14 days after second dose). Among partially immunized (≥14 days after first dose and before second dose) persons, it was reported to be 0.19 infections per 1,000 person-days.

While most studies so far have reported on impact of vaccination among the common public, the present study gives exclusivity to patients diagnosed with COVID-19 comparing the cost/benefit between vaccinated and un-vaccinated patients. Among the COVID-19 patients included in our study 86% had not received vaccination. Among those who provided reasons for not vaccinating themselves, in the vast majority the reasons were trivial ones like apprehension/fear, lack of interest, ignorance, low confidence in vaccines due to misinformation, disinformation, rumors, and conspiracy theories, in particular through social media. WHO listed vaccine hesitancy among the top 10 global threats to health in 2019. One recent study showed that vaccine hesitancy is common and is due to concerns about unknown future effects, side effects, and a lack of trust [11]. Our study also shows that an active demystification program and campaign through community engagement will make more people voluntarily recruit themselves for vaccination.

Among those who contracted COVID-19, post vaccination there was significant reduction in Average length of stay (ALOS), ICU stay and thus the total cost of treatment compared to the un-vaccinated group. There was 66% risk ratio reduction of ICU stay among those who have completed two doses of vaccination. Even among those with comorbidities, a single dose was able to significantly reduce ALOS and hospital expenses. Studies [12] have shown 60-70% reduction in infection risk starting 12 days after the first dose. Tyagi et al [4] in a survey among health care workers, with a higher viral load, reported 17% break through infections post vaccination, although all of them were mild. Studies [13] have shown that a single dose of vaccine was 75 to 80% effective while two doses were 95-99% effective in reducing mortality among COVID-19 patients. Our study has also shown that there was 81% relative risk reduction in mortality by vaccination.

Another important public health challenge, during vaccine shortage, is whether to continue vaccinating public for first dose or complete the second dose earlier. A recent study conducted by University College London[14] revealed that more than 90 per cent of Britons produced antibodies to COVID-19 after having a single dose of vaccine. The study found that 96% of people, who had the vaccine, had developed antibodies 28 days after their first dose. Romero[5] in a simulation based modelling study showed that delaying second vaccine doses for people younger than 65 to prioritize people getting their first dose could reduce mortality. Our findings show that there was a significant reduction in hospital expenses and ICU requirement after a first dose vaccination. Completion of two doses was associated with further reduction in hospital expenses, ICU requirement and mortality. A recent UK study [16] also stressed the importance of vaccinating the entire population with two doses quickly so that the country develops herd immunity.

There are several limitations in this study. Being a retrospective cohort study, in analyzing vaccination status among COVID-19 patients, we were unable to comment on the incidence of disease among the vaccinated/ unvaccinated population. This study covers patients under health insurance cover who are treated in private settings, thus excluding a larger population treated at government hospitals. However it covers pan-India data and is likely to represent a national trend. Further this study focused only on those aged 45+ and happened during the early phase of vaccination when skepticism was high and vaccine penetration was low.

This study included only hospitalized patients, while a larger proportion of milder cases post vaccination could have been treated as outpatient or at home care settings. This study also did not cover asymptomatic cases that could have happened post vaccination. While vaccination is known to reduce symptomatic disease, hospitalization and death, Tang et al [15] in a study showed that vaccination also reduced asymptomatic COVID-19. Further larger prospective studies are needed to see whether vaccination would also reduce transmissibility of the disease.

In our study we did not have details of the strain that could have been responsible for the break through infection. Also it was difficult to assess the baseline immune status of the population that could have attributed to this. A recent study[16] from UK showed that 33% protection was noted after a single dose of vaccination against B 1.617.2 variant, while this increased to 60% two weeks after second dose. Future research should focus on the efficacy of vaccines against the specific variants of concern.

## Conclusions

Our study demonstrates a significant reduction in average length of stay, ICU requirement, mortality and the total cost of treatment for patients who have completed two doses of vaccination. Among those who received two doses of vaccination there was a 66% relative risk reduction in ICU requirement and 81% relative risk reduction in mortality. The current study covering a pan-India data from a health insurance point of view is one of its kind to analyze not only the medical benefits but also the financial implications. This may pave way in educating and motivating the public regarding the role of vaccination in reducing morbidity, mortality and hospital expenses. This in turn is likely to promote vaccination drive in a big way among rural and urban population alike.

## Data Availability

Data may be available on request (without patient identity)

## Highlights

- Vaccine hesitancy is common in the Indian subcontinent essentially due to fear, myths and misinformation.
- Our study demonstrates a significant reduction in average length of stay, ICU requirement, mortality and the total cost of treatment for patients who have completed two doses of vaccination.
- Among those who received two doses of vaccination there was a 66% relative risk reduction in ICU requirement and 81% relative risk reduction in mortality. These findings may be used in motivating public and promoting vaccination drive.

